# Reproducibility and associated regression dilution bias of accelerometer-derived physical activity and sleep in the UK Biobank

**DOI:** 10.1101/2025.01.16.25320679

**Authors:** Charilaos Zisou, Hannah Taylor, Ben Lacey, Imen Hammami, Rosemary Walmsley, Tessa Strain, Katrien Wijndaele, Karl Smith-Byrne, Derrick Bennett, Sarah Lewington, Jemma C. Hopewell, Aiden Doherty

## Abstract

**Background:** Previous studies on the reproducibility of 7-day accelerometer measurements have been limited by small sample sizes and short follow-up periods. We aimed to assess the long-term reproducibility of accelerometer-derived physical activity and sleep, and to illustrate the impact of regression dilution bias on the association between daily step count and coronary heart disease (CHD) in UK Biobank (UKB).

**Methods:** We analysed data from 3138 UKB participants in the main accelerometry sub-study with up to four repeat accelerometer measurements after 3-4 years. Nine physical activity and sleep phenotypes were extracted to capture different movement behaviours. Reproducibility was assessed using intraclass correlation coefficients (ICCs). The impact on disease associations was illustrated by considering daily step count and incident CHD using Cox regression (87 180 participants; 3899 CHD events), before and after correction for regression dilution.

**Results:** Among the 3138 participants, 51% were women and the mean (SD) age was 63.1 (9.4) years. Reproducibility of phenotypes was moderate to good, with the ICC (95% CI) for overall activity at 0.75 (0.74-0.76), and individual phenotypes ranging from 0.58 (0.56-0.59) for sleep efficiency to 0.69 (0.68-0.70) for sedentary behaviour. In our example, the inverse association between daily step count and CHD showed a 20% lower risk of CHD per 4000 usual steps after correcting for regression dilution, compared to 13% before correction.

**Conclusions:** Accelerometer measurements are moderately reproducible and comparable to measures like blood pressure. Correcting for regression dilution bias is crucial to quantify associations of usual physical activity and sleep with disease risk.

**Key Messages:**

- What was your research question?

- Accelerometer-derived physical activity and sleep are increasingly used in large cohort studies, but the long-term reproducibility of these measures and the extent to which their associations with health outcomes are impacted by regression dilution bias remain poorly understood.
- What did you find?

- In this study of repeat 7-day accelerometer measurements in 3138 UK adults, the agreement between measurements taken 3.7 years apart ranged from 58% for sleep efficiency to 75% for overall activity.
- Consideration of regression dilution in an example association between daily step count and incident coronary heart disease illustrated how associations of accelerometer measures with disease risk have been substantially underestimated.
- Why is it important?

- Incorporation of repeat accelerometer measurements in cohort studies will allow researchers to quantify their reproducibility and correct epidemiological associations for regression dilution bias.

## Introduction

Physical inactivity is associated with several common diseases, including cardiovascular disease and type 2 diabetes.^1^ Variations in sleep behaviour are also linked to adverse health outcomes, such as cardiometabolic and mental health conditions.^2^ To better understand these relationships, large cohort studies have increasingly incorporated device-based measurements of movement behaviours using wearable accelerometers.^3-5^ These measurements, which are less prone to information biases than self-reported questionnaires and can track movements continuously throughout the day, have provided novel insights into the health effects of physical activity and sleep. For example, the inverse associations between device-measured physical activity and cardiovascular and all-cause mortality are much stronger than previously observed from self-reported data.^6^ Moreover, device-based studies challenge prior findings that long sleep duration is associated with worse health outcomes.^7,8^

Previous study designs have typically instructed participants to wear an accelerometer for seven consecutive days.^9^ This approach balances the need for extensive data collection with logistical challenges, such as device battery life and participant compliance. However, the long-term reproducibility of 7-day accelerometer measurements (i.e., the degree of agreement of repeat measurements within the same participants over time) remains uncertain, largely due to previous studies having small sample sizes and collecting repeat measurements over short intervals.^10-14^ Within-person variation from measurement error, short-term fluctuations and longer-term changes in activity patterns may lead to measurements that do not accurately reflect an individual’s ‘usual’ or habitual movement behaviours over the long term. This variation systematically weakens observed associations with health outcomes, an effect known as regression dilution bias.^15^ Estimating the reproducibility of accelerometer-derived phenotypes enables correction for this bias, providing more accurate estimates of associations between usual movement behaviours and health outcomes.^16^ Although correction methods have been commonly applied to other exposures such as blood pressure and total cholesterol,^15,17,18^ they have rarely been utilised in analyses of movement behaviours,^19^ especially with device-based data.

This study aimed to 1) investigate the long-term reproducibility of accelerometer-derived phenotypes of physical activity and sleep using repeat 7-day accelerometer measurements, and 2) illustrate the impact of regression dilution bias through an example examining the association between daily step count and incident coronary heart disease (CHD).

## Methods

### Study design and population

Details of the UK Biobank study design have been extensively reported elsewhere.^20^ Briefly, between 2006-2010, 0.5 million participants aged 40-69 years from England, Scotland and Wales were recruited in a prospective cohort study (response rate 5.5%). During a baseline visit to an assessment centre, participants completed touchscreen questionnaires, were interviewed by trained nurses and had anthropometric measurements and biological samples taken.^21^ A subset of 103 712 individuals also participated in the main accelerometry sub-study between 2013-2015.^4^ Participants were sent an Axivity AX3 triaxial accelerometer (100Hz) by post and asked to wear it continuously on their dominant wrist for seven days during normal activities. At the end of the recording period, participants were asked to mail the device back to the coordinating centre.

Between 2017-2019, a sample of 5067 participants from the main accelerometry sub-study with good accelerometer compliance (near-complete 7-day wear and prompt device return) were invited by email to undergo repeat accelerometer measurements (**Figure 1**). This sample was selected using a stratified random sampling procedure to ensure equal representation across sex and age groups (10-year intervals). Participants were asked to provide four measurements spaced approximately 3 months apart, allowing for some participants to miss initial measurements but complete subsequent ones. Of those invited, 3208 (63%) accepted the invitation. Devices were dispatched seasonally over four cycles: November 2017–May 2018 (3202 devices dispatched, 2634 measurements released), February–August 2018 (3125 devices dispatched, 3095 measurements released), May–November 2018 (3070 devices dispatched, 3039 measurements released), and August 2018–February 2019 (3051 devices dispatched, 3012 measurements released). There were minor differences between devices returned and data released, largely due to issues with data reading; however, the first cycle was additionally subject to device malfunctions in 17% of devices distributed. Overall, there were 38 participants with only one measurement, 91 with two measurements, 716 with three measurements and 2353 participants with all four measurements. Data collection followed the same protocol as the main accelerometry sub-study.

**Figure 1:**
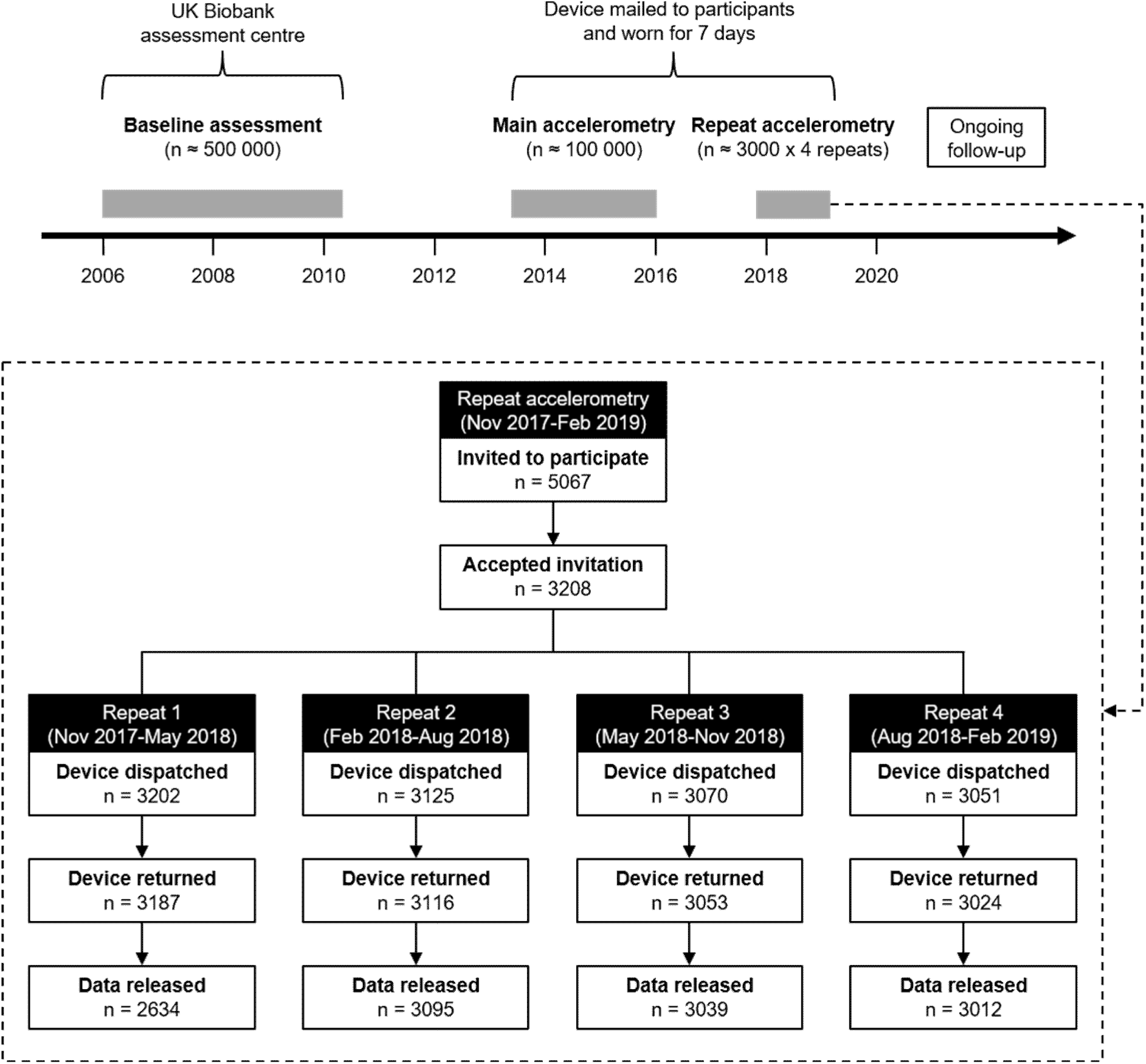
Timeline of UK Biobank study assessments and flow diagram of repeat accelerometry sub-study participants. Differences in devices dispatched among repeat cycles are largely due to participants opting out. Differences between devices returned and data released within each repeat cycle are largely due to issues with reading accelerometer data; however, the first cycle was additionally subject to device malfunction in 17% of devices distributed.

### Accelerometer-derived phenotypes of physical activity and sleep

The raw accelerometer time series were pre-processed using established methods and the average overall activity was extracted.^4^ Validated machine learning models were used to classify movement behaviours at the 30-second level and derive summary physical activity and sleep phenotypes, averaged over the week to provide daily values. A self-supervised neural network and a peak detection algorithm were used to measure step count, defined as the median steps per day (https://github.com/OxWearables/stepcount).^22^ Additionally, peak 30-minute cadence was calculated as the average steps per minute from the 30 most active (not necessarily consecutive) minutes of the day. Daily durations of moderate-to-vigorous physical activity (MVPA), light physical activity, sedentary behaviour, and time spent in bed were estimated using a random forest and hidden Markov model (https://github.com/OxWearables/biobankAccelerometerAnalysis).^23^ Overnight sleep duration, defined as the median hours of sleep per night, and sleep efficiency, calculated as the ratio of overnight sleep duration to total time spent in bed, were measured using a self-supervised neural network (https://github.com/OxWearables/asleep).^7^

### Ascertainment of coronary heart disease

Participants were followed up from the end date of the main accelerometer wear through electronic health record linkage to UK hospital inpatient and operations data and the UK death register to obtain incident CHD events (**Table S1**). Participants without CHD recorded were censored at the end of linkage data availability (31 October 2022, 31 August 2022 and 31 May 2022 for participants in England, Scotland and Wales, respectively), loss to follow-up, or death, whichever occurred first.

### Statistical analysis

Participants with low-quality accelerometer data, namely those whose device could not be processed or calibrated, who had unrealistically high acceleration values (>100mg) or who had insufficient wear time (<3 days of wear or missing data for the same hour across all recorded days in the week), were excluded.^4^ For sleep phenotypes, we additionally excluded days with less than 22 hours of wear time.^7^ Only data meeting these quality criteria are hereby referred to as “valid data”. Accelerometer phenotypes were compared across the main and the repeat measurements using descriptive statistics; continuous variables were approximately normally distributed and were expressed as mean (standard deviation [SD]), except for MVPA and sleep efficiency, which were skewed and were described as median (interquartile range [IQR]).

To assess reproducibility, participants were divided into fifths based on their main accelerometer measurements and changes in mean phenotypes within those fifths were tracked after 4 years. Some participants may have had unusually active or inactive weeks when initially measured, but calculating the group means of the repeat measurements yields unbiased estimates of usual group levels at the time of measurement.^15^ Additionally, intraclass correlation coefficients (ICCs) were calculated, which assess reproducibility and can also be used to correct for regression dilution bias in epidemiological association studies.^16^ ICCs and 95% confidence intervals (CIs) were estimated using a two-way mixed effects model with absolute agreement.^24^ Statistical information was incorporated from all measurements by calculating the ICCs between the main and each of the repeat measurements and obtaining their inverse-variance weighted average. To account for seasonal variations, all measurements were first regressed on the corresponding season of accelerometer wear and the residuals of that regression were used to calculate the ICCs. For skewed phenotypes (MVPA and sleep efficiency), ICCs were assessed with and without inverse-normal transformations using a rank-based approach. Stratified analyses were conducted to assess ICCs by subgroups of age at main accelerometry sub-study entry (<60, ≥60 years), sex, body mass index (<25, ≥25 kg/m^2^) and prior self-reported illness or disability (healthy or prior disease). Sensitivity analyses were performed by restricting the dataset to participants with four valid repeat measurements to assess whether variations in measurements across cycles were due to differences in participant characteristics.

Regression dilution bias, resulting from measurement error, short- and long-term changes in activity levels, leads to a systematic underestimation of observed epidemiological associations. To illustrate this, we considered the association between daily step count and CHD as an example, which has been consistently observed in previous studies.^25,26^ Using all 103 712 participants from the UKB main accelerometry sub-study, we excluded those who withdrew from the study, had low-quality accelerometer data (quality criteria described above), had atherosclerotic cardiovascular disease prior to the main accelerometer wear (to limit reverse causation), or had missing covariates. Cox regression models were used to estimate hazard ratios (HRs) and 95% CIs for CHD, with step count analysed both across fifths and as a continuous variable (per 4000 steps, which is approximately equal to the SD), adjusting for potential confounders (**Supplementary Methods**; **Table S2**). To assess reverse causation bias, a sensitivity analysis was conducted by excluding CHD events occurring within the first 2 years of follow-up, as done in previous studies.^23,26^

To illustrate the correction for regression dilution bias visually, we plotted the HR of each step count fifth defined by the main accelerometer measurements against the inverse-variance weighted average over the repeat measurements, estimating each fifth’s usual daily step count.^15^ The log HR per 4000 steps and its standard error were divided by the corresponding ICC to obtain the HR per 4000 usual steps.^16^ A sensitivity analysis assessed the difference in estimates when age- and sex-specific corrections for regression dilution bias were used (**Supplementary Methods**).

The processing of accelerometer data was undertaken in Python (version 3.9) and all statistical analyses were performed in R (version 4.3).

## Results

### Reproducibility analysis - Participant characteristics

After quality-related exclusions, 3138 participants with valid accelerometer data from the main sub-study and at least one valid repeat measurement were included in the reproducibility analysis (**Table 1**; **Figure S1**). Mean (SD) age was 63.1 (9.4) years, 1613 (51.4%) were women and 3038 (96.8%) identified as White. Compared to participants in the main accelerometry sub-study, those with repeat measurements were younger at baseline (53.8 vs. 56.7 years) and less likely to be women (51.4% vs. 56.3%). Additionally, repeat accelerometry participants were more likely to have a university degree and less likely to be taking blood pressure and cholesterol medication or have prevalent cardiovascular disease. Other characteristics were broadly similar between the main and repeat accelerometry sub-studies. However, compared to the entire UK Biobank cohort, the accelerometry participants generally exhibited higher socio-economic status, lower rates of smoking, a lower body mass index and a lower prevalence of health conditions and medications, potentially indicative of a healthy volunteer effect.^27^

**Table 1:** Selected baseline characteristics of all UK Biobank participants, those who participated in the main accelerometry sub-study and those who had at least one valid repeat accelerometer measurement.

| Characteristic at baseline assessment | Baseline assessment<br>2006-2010<br>(n=502 364) | Main accelerometry<br>2013-2015<br>(n=95 981) | Repeat accelerometry<br>2017-2019<br>(n=3138) |
| --- | --- | --- | --- |
| Demographic and socio-economic factors |  |  |  |
| Age at baseline, years | 57.0 (8.1) | 56.7 (7.8) | 53.8 (9.3) |
| Current age*, years | 57.0 (8.1) | 62.4 (7.8) | 63.1 (9.4) |
| Women | 273 297 (54.4%) | 54 038 (56.3%) | 1613 (51.4%) |
| White | 472 569 (94.1%) | 92 687 (96.6%) | 3038 (96.8%) |
| College or University degree | 161 102 (32.1%) | 41 289 (43.0%) | 1476 (47.0%) |
| Townsend deprivation index | -2.1 (-3.6, 0.5) | -2.4 (-3.8, -0.2) | -2.3 (-3.7, 0) |
| Lifestyle factors |  |  |  |
| Current smoker | 52 960 (10.5%) | 6613 (6.9%) | 195 (6.2%) |
| Daily alcohol drinker | 101 745 (20.3%) | 21 966 (22.9%) | 681 (21.7%) |
| Fruits and vegetables $\geq 8$ servings/day | 183 677 (36.6%) | 35 557 (37.0%) | 1091 (34.8%) |
| Physical and blood measurements |  |  |  |
| Body mass index, kg/m<sup>2</sup> | 27.4 (4.8) | 26.7 (4.5) | 26.4 (4.6) |
| Systolic blood pressure, mmHg | 137.9 (18.6) | 136.5 (18.2) | 134.9 (17.8) |
| Resting heart rate, bpm | 69.4 (11.3) | 68.5 (10.9) | 68.3 (11.0) |
| LDL cholesterol, mmol/L | 3.6 (0.9) | 3.6 (0.8) | 3.5 (0.8) |
| Self-reported conditions and medication |  |  |  |
| Blood pressure medication | 103 982 (20.7%) | 16 240 (16.9%) | 408 (13.0%) |
| Cholesterol medication | 86 874 (17.3%) | 13 475 (14.0%) | 350 (11.2%) |
| Long-standing illness or disability | 159 853 (31.8%) | 26 762 (27.9%) | 827 (26.4%) |
| Poor overall health | 22 768 (4.5%) | 2397 (2.5%) | 77 (2.5%) |
| Atherosclerotic cardiovascular disease** | 37 030 (7.4%) | 7327 (7.6%) | 186 (5.9%) |

Mean (SD) or median (IQR) shown for continuous variables. N (%) shown for categorical variables. Characteristics were measured during the baseline assessment (2006-2010), unless otherwise stated. Participants with low-quality accelerometer data were excluded. \*Refers to participants' ages at the time of each assessment. \*\*Diagnosed prior to each assessment, as recorded in hospital records or self-reported.

The repeat measurements were conducted 3.2 to 4.0 years (mean [SD] of 3.6 [0.7]) after the main accelerometry sub-study (**Table 2**). Participants in all cycles had the same mean age at the main sub-study, and the proportion of women remained consistent. Data were primarily collected in winter (70%) during cycle 1, spring (78%) during cycle 2, summer (78%) during cycle 3 and autumn (79%) during cycle 4. Mean levels of physical activity and sleep were similar across the main and repeat measurement cycles for all phenotypes, with small seasonal differences.

**Table 2:**
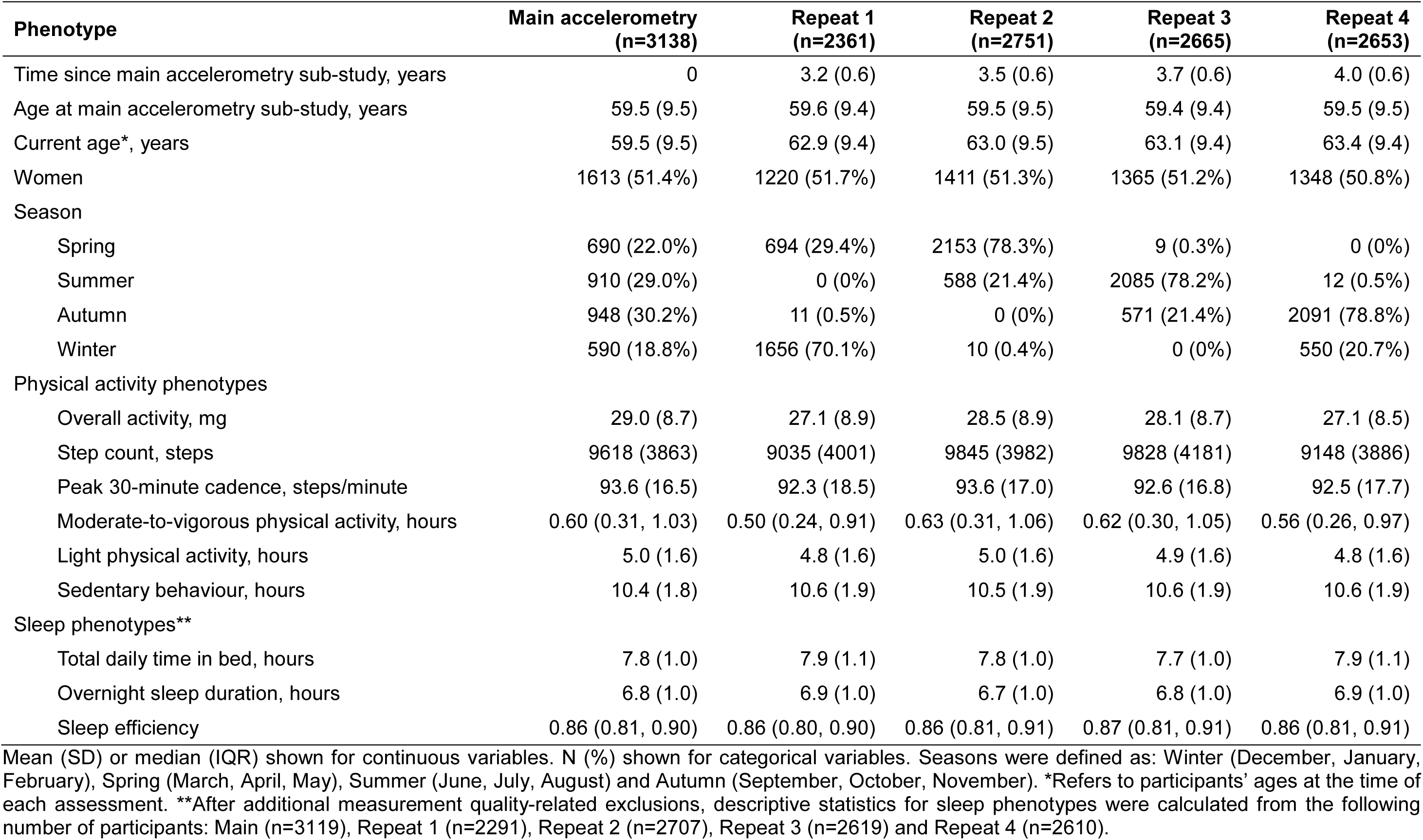
Accelerometer-derived phenotypes of physical activity and sleep across main and repeat accelerometry sub-studies.

| Phenotype | Main accelerometry<br>(n=3138) | Repeat 1<br>(n=2361) | Repeat 2<br>(n=2751) | Repeat 3<br>(n=2665) | Repeat 4<br>(n=2653) |
| --- | --- | --- | --- | --- | --- |
| Time since main accelerometry sub-study, years | 0 | 3.2 (0.6) | 3.5 (0.6) | 3.7 (0.6) | 4.0 (0.6) |
| Age at main accelerometry sub-study, years | 59.5 (9.5) | 59.6 (9.4) | 59.5 (9.5) | 59.4 (9.4) | 59.5 (9.5) |
| Current age*, years | 59.5 (9.5) | 62.9 (9.4) | 63.0 (9.5) | 63.1 (9.4) | 63.4 (9.4) |
| Women | 1613 (51.4%) | 1220 (51.7%) | 1411 (51.3%) | 1365 (51.2%) | 1348 (50.8%) |
| Season |  |  |  |  |  |
| Spring | 690 (22.0%) | 694 (29.4%) | 2153 (78.3%) | 9 (0.3%) | 0 (0%) |
| Summer | 910 (29.0%) | 0 (0%) | 588 (21.4%) | 2085 (78.2%) | 12 (0.5%) |
| Autumn | 948 (30.2%) | 11 (0.5%) | 0 (0%) | 571 (21.4%) | 2091 (78.8%) |
| Winter | 590 (18.8%) | 1656 (70.1%) | 10 (0.4%) | 0 (0%) | 550 (20.7%) |
| Physical activity phenotypes |  |  |  |  |  |
| Overall activity, mg | 29.0 (8.7) | 27.1 (8.9) | 28.5 (8.9) | 28.1 (8.7) | 27.1 (8.5) |
| Step count, steps | 9618 (3863) | 9035 (4001) | 9845 (3982) | 9828 (4181) | 9148 (3886) |
| Peak 30-minute cadence, steps/minute | 93.6 (16.5) | 92.3 (18.5) | 93.6 (17.0) | 92.6 (16.8) | 92.5 (17.7) |
| Moderate-to-vigorous physical activity, hours | 0.60 (0.31, 1.03) | 0.50 (0.24, 0.91) | 0.63 (0.31, 1.06) | 0.62 (0.30, 1.05) | 0.56 (0.26, 0.97) |
| Light physical activity, hours | 5.0 (1.6) | 4.8 (1.6) | 5.0 (1.6) | 4.9 (1.6) | 4.8 (1.6) |
| Sedentary behaviour, hours | 10.4 (1.8) | 10.6 (1.9) | 10.5 (1.9) | 10.6 (1.9) | 10.6 (1.9) |
| Sleep phenotypes** |  |  |  |  |  |
| Total daily time in bed, hours | 7.8 (1.0) | 7.9 (1.1) | 7.8 (1.0) | 7.7 (1.0) | 7.9 (1.1) |
| Overnight sleep duration, hours | 6.8 (1.0) | 6.9 (1.0) | 6.7 (1.0) | 6.8 (1.0) | 6.9 (1.0) |
| Sleep efficiency | 0.86 (0.81, 0.90) | 0.86 (0.80, 0.90) | 0.86 (0.81, 0.91) | 0.87 (0.81, 0.91) | 0.86 (0.81, 0.91) |

Mean (SD) or median (IQR) shown for continuous variables. N (%) shown for categorical variables. Seasons were defined as: Winter (December, January, February), Spring (March, April, May), Summer (June, July, August) and Autumn (September, October, November). \*Refers to participants' ages at the time of each assessment. \*\*After additional measurement quality-related exclusions, descriptive statistics for sleep phenotypes were calculated from the following number of participants: Main (n=3119), Repeat 1 (n=2291), Repeat 2 (n=2707), Repeat 3 (n=2619) and Repeat 4 (n=2610).

### Reproducibility of accelerometer-derived phenotypes of physical activity and sleep

The mean values of all phenotypes within the fifths defined by the main measurements moderately converged after 4 years, displaying a pattern of regression towards the mean with small seasonal variations (**Figure 2**). The top fifth converged more than the bottom fifth for physical activity phenotypes, while the opposite trend was observed for sedentary behaviour and sleep phenotypes. The reproducibility of the examined phenotypes was moderate to good,^24^ with the ICC (95% CI) for overall activity being 0.75 (0.74-0.76) and the rest ranging from 0.58 (0.56-0.59) for sleep efficiency to 0.69 (0.68-0.70) for sedentary behaviour (**Figure 2**; **Table S3**). There was no material difference between the ICCs with and without inverse-normal transformations; therefore, only the latter are presented. Reproducibility was generally higher for older (≥60 years) compared to younger (<60 years) individuals and for women compared to men (**Table S4**). Minor differences in reproducibility were observed across some physical activity phenotypes between normal/underweight (<25 kg/m^2^) and overweight/obese individuals (≥25 kg/m^2^; **Table S5**). Individuals with prior illness or disability exhibited slightly higher reproducibility compared to healthy individuals, with step count showing a more pronounced difference. Restricting the analysis to participants with four valid repeat measurements did not substantially alter the results (**Tables S6**, **S7**).

**Figure 2:**
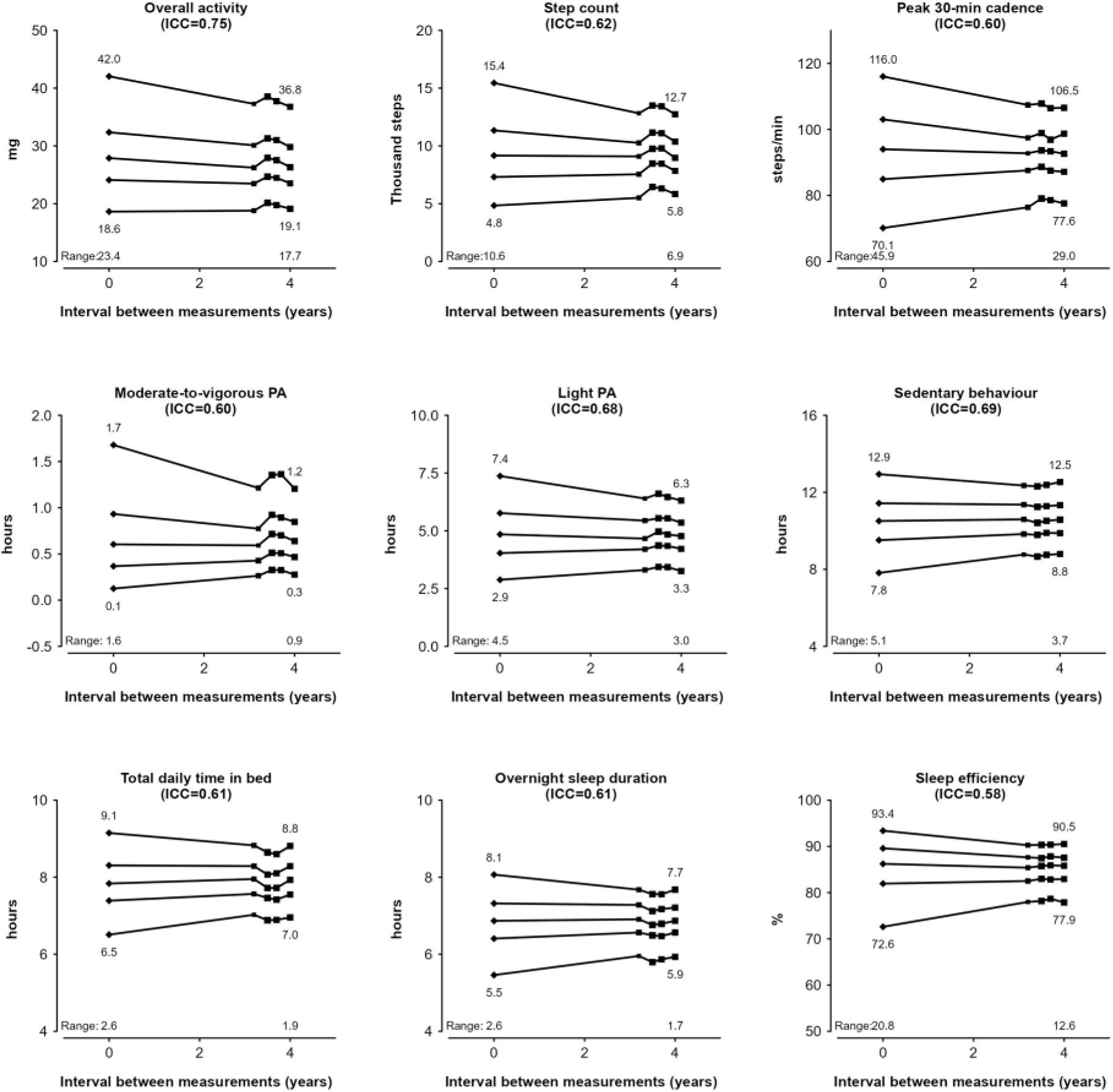
Changes in accelerometer-derived phenotypes of physical activity and sleep within fifths defined by the main accelerometer measurements. Analysis was done in 3138 participants with valid accelerometer data from the main sub-study and at least one valid repeat measurement. Squares represent mean values initially and at subsequent repeat measurements for participants divided into fifths according to their main accelerometer measurement. The size of each square is proportional to the number of participants available. Mean values in the top and bottom groups and the absolute differences (ranges) between them are given at years 0 and 4. ICC, Intraclass correlation coefficient; PA, Physical activity.

### Illustrative example: Impact of regression dilution bias on the association between daily step count and CHD

Of the 103 712 participants with accelerometer data, 87 180 remained in the disease association analysis after excluding those who withdrew from the study (n=52), had low-quality accelerometer data (n=7679), had atherosclerotic cardiovascular disease prior to the main accelerometer wear (n=7327) or had missing covariates (n=1474; **Figure S2**). During a median (IQR) follow-up of 7.9 (7.3-8.4) years, 3899 incident CHD events occurred. Every additional 4000 steps per day was associated with a 13% lower risk of CHD (HR 0.87, 95% CI 0.84-0.90) after adjusting for potential confounders with no correction for regression dilution bias (**Figure 3**). Plotting the estimated HRs against the inverse-variance weighted group means at the repeat measurements revealed a substantially stronger association between usual daily step count and CHD. After correcting for regression dilution bias (by dividing the logHR and standard error by the corresponding ICC of 0.62), every additional 4000 usual steps per day was associated with a 20% lower risk of CHD (HR 0.80, 95% CI 0.76-0.85; excluding the first 2 years of follow-up did not materially alter the estimate; **Table S8**). Sensitivity analysis using age- and sex-specific corrections for regression dilution bias resulted in highly consistent estimates (**Table S9**).

**Figure 3:**
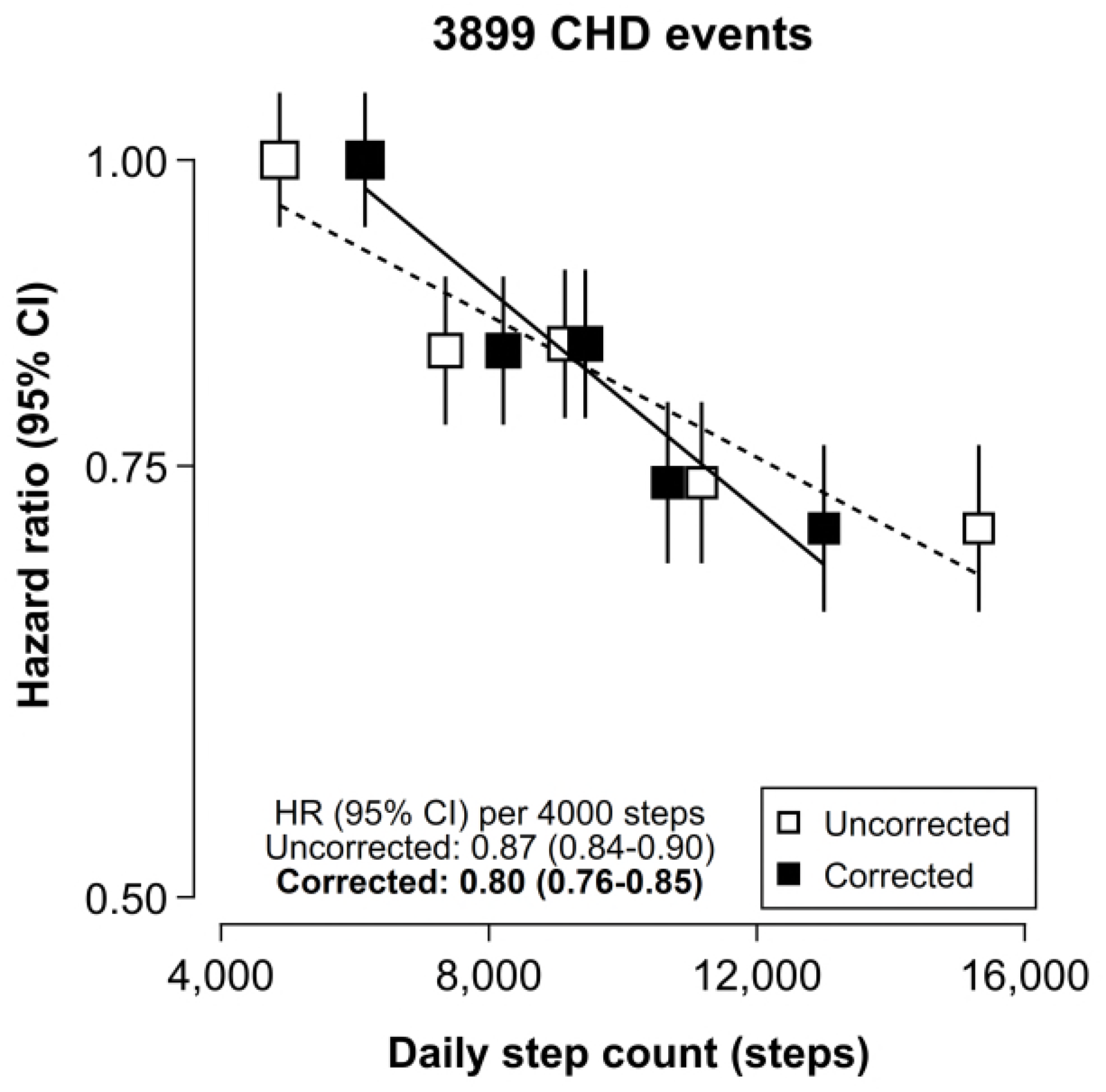
Illustrative example of the association between daily step count and incident coronary heart disease, before and after correction for regression dilution bias. Analysis was done in 87 180 participants after excluding those with low-quality accelerometer data, prevalent atherosclerotic cardiovascular disease, or missing covariates. Cox models used age as the time scale, were stratified by sex and adjusted for season of accelerometer wear, ethnicity, geographical area, education, Townsend deprivation index, alcohol intake, smoking status, and fresh fruit and vegetable intake. Squares represent hazard ratios (HRs) with their size being inversely proportional to the group-specific variance of the log HR. Vertical lines represent group-specific 95% confidence intervals (CIs). Diagonal lines correspond to slopes of weighted linear regressions with weights equal to the inverse variance of the log HRs. CHD, Coronary heart disease, HR, Hazard ratio.

## Discussion

In the largest study of repeat 7-day accelerometer measurements to date, mean levels of physical activity and sleep within groups defined by their initial accelerometer measurements moderately converged after 4 years, indicating some regression towards the mean. Reproducibility was moderate to good, with overall activity reaching 75%, and individual phenotypes ranging from 58% for sleep efficiency to 69% for sedentary behaviour. Reproducibility was generally higher in women compared to men, and in older compared to younger individuals. Furthermore, correction for regression dilution bias was shown to have a material impact on epidemiological associations, underestimating effects by nearly 40% in our example of daily step count and CHD.

Our reproducibility results are broadly consistent with previous smaller studies using hip-worn accelerometers, despite differences in study design. For overall activity, reproducibility estimates have been reported between 76-83%.^10,13,14^ Furthermore, a study of 1679 adults reported a correlation of 55% between repeat measurements of step counts taken approximately 3.7 years apart, which closely aligns with our estimate.^28^ For MVPA, light physical activity and sedentary behaviour, reproducibility estimates show a greater degree of heterogeneity, between 59-77%.^10,12-14^ This heterogeneity is possibly driven by differences in study timeframes and population characteristics, particularly age and sex distributions. For sleep phenotypes, a study with repeat wrist-worn accelerometer measurements over a 1-year period reported correlations of 67% for total time in bed, 76% for overnight sleep duration, and an unexpectedly high 90% for sleep efficiency, though these measurements relied on participant input.^29^ In contrast to these smaller studies, our analysis identified clear differences in reproducibility by age, sex, and prior disability status, offering important insights into subgroup-specific variability.

Compared to self-report, accelerometer-derived physical activity and sleep exhibit markedly higher reproducibility, further highlighting the value of incorporating device-based measurements in large-scale epidemiological studies. A study of 19 000 UK Biobank participants reported the reproducibility of self-reported total physical activity at around 50% after 4.3 years, with estimates for different movement behaviours ranging from 45% to 60%.^30^ Similar results were obtained in a Chinese population of 20 000 participants over a 3-year period.^19^ For self-reported sleep, two studies in UK populations (n=13 000 over 1.7 years, and n=19 000 over 4.3 years) found kappa scores around 50% for sleep duration categories.^30,31^ Notably, the reproducibility of accelerometer-derived phenotypes is comparable to that of well-established CHD risk factors like systolic blood pressure and total cholesterol, which is 65-70% within a 5-year period.^17,18^ In the broader context of epidemiological research, accelerometer-derived phenotypes exhibit moderately lower reproducibility than the most reproducible imaging measures (median=85%) and substantially higher reproducibility than the least reproducible 24-hour dietary recall measures collected through online questionnaires (median=35%), based on a UK Biobank study of 2858 variables.^32^

The reproducibility of accelerometer measurements is sufficiently high for use in epidemiological studies; however, measurement error and fluctuations in physical activity and sleep still occur. These deviations from an individual’s usual physical activity and sleep levels lead to underestimations of associations with health outcomes in studies based on single assessments, as illustrated by our example of daily step count and CHD. Our example focuses on the relative change in risk after correction for regression dilution bias. However, for epidemiological studies focusing on disease associations, comprehensive consideration of other sources of bias (e.g., residual confounding and reverse causation) along with regression dilution bias, is important for appropriate interpretation. Few large studies of physical activity and sleep relying solely on self-report have employed repeat assessments. For example, studies using averages of repeated surveys to characterise physical activity levels have found stronger associations with health outcomes, suggesting prior underestimations by around 60%.^33,34^ The approach used in our study has the advantage that only a subset of the original cohort needs to be resurveyed, a more efficient method that has successfully been applied in UK and Chinese population cohorts.^19,31^

This study has several strengths, including being the largest reproducibility study of device-measured physical activity and sleep to date, with four repeat measurements, which enabled us to assess reproducibility across key subgroups. Additionally, it focused on wrist-worn accelerometers, which are increasingly used in large-scale cohort studies, and examined a broad range of derived phenotypes, capturing different aspects of movement behaviour. However, the study also has limitations. First, participant characteristics were measured at baseline, a few years prior to the main accelerometer measurements. As a result, these characteristics (e.g., body mass index and self-reported disability used in the stratified analyses) may not accurately reflect participants’ characteristics at the time of the accelerometer measurements. Second, the UK Biobank is not representative of the UK population, so our ICCs may not generalise to populations with different movement behaviour patterns, though our estimates align broadly with those from other studies. Finally, incorporating methods to account for measurement error in the confounders could further strengthen epidemiological association estimates in future studies.^35^

## Conclusion

This study provides new insights into the moderate to good long-term reproducibility of accelerometer-derived physical activity and sleep among UK adults. Similarly to other measures, such as blood pressure, this level of reproducibility is sufficiently high for epidemiological research, but associations with health outcomes remain subject to regression dilution bias. This highlights the need for cohorts to incorporate repeat accelerometer measurements, and for researchers to account for regression dilution bias in epidemiological studies of accelerometer-derived physical activity and sleep.

## Ethics approval

Ethics approval for this study is covered by the general ethics approval for UK Biobank studies from the North West Multi-centre Research Ethics Committee (Ref 11/NW/0382), which is renewed every five years. All study participants provided written informed consent.

## Data availability

Researchers can request data access by following the UK Biobank’s data access guidelines available at (https://www.ukbiobank.ac.uk/enable-your-research).

## Author contributions

CZ, DB, SL, JCH and AD conceived the project and developed an analysis plan. CZ performed all analyses and wrote the first draft of the manuscript. JCH and AD supervised the work. All the authors discussed the results and contributed to the final manuscript.

## Acknowledgements

We thank Dr. Catherine Calvin, Howard Callen, and Dr. Janet Maccora from the UK Biobank team for providing key information and guidance on the study design of the repeat accelerometry sub-study. This research was conducted using the UK Biobank resource under application number 59070. We are grateful to all participants for generously contributing their data to advance research in population health. This work used data provided by patients and collected by the NHS as part of their care and support. Computation used the Oxford Biomedical Research Computing (BMRC) facility, a joint development between the Wellcome Centre for Human Genetics and the Big Data Institute supported by Health Data Research UK and the NIHR Oxford Biomedical Research Centre. The views expressed are those of the authors and not necessarily those of the NHS, the NIHR or the Department of Health.

