## Supplementary material for "Reproducibility and associated regression dilution bias of accelerometer-derived physical activity and sleep in the UK Biobank"

**Table S5:** Intraclass correlation coefficients (95% CIs) of accelerometer-derived phenotypes of physical activity and sleep, by subgroups of body mass index and self-reported disability11

### Supplementary Methods

*Association between daily step count and incident coronary heart disease (CHD)*

Participants were divided into fifths based on their step counts from the main accelerometry sub-study and Cox regression models were used to estimate hazard ratios (HRs) and 95% CIs for CHD in each fifth. Age was used as the time scale and the models were stratified by sex and adjusted for season of accelerometer wear (winter, spring, summer, autumn; included as a technical covariate), ethnicity (Asian, Black, White, other), geographical region (10 regions), education (none, national exams at ages 16-18 years, vocational qualifications, college or university degree), Townsend deprivation index (quintiles), alcohol intake frequency (<1 times/week, 1-2 times/week, 3-4 times/week, daily), smoking status (never, former, current), and fresh fruit and vegetable intake (<4, 4-5.9, 6-7.9, ≥8 servings/day; **Table S2**). CIs were calculated using the group-specific variances of the log HRs (including that of the reference group), thus allowing comparisons between any two groups.[1] Additionally, step count was modelled in continuous form to calculate the HR for CHD per 4000 steps (approximately equal to the SD). The proportional hazards assumptions for the Cox models were assessed by examining the plots of the Schoenfeld residuals and were satisfied.[2]

*Age- and sex-specific correction for regression dilution bias*

A sensitivity analysis assessed the difference in the estimates when age- and sex-specific corrections for regression dilution bias were used. The log HRs per 4000 steps were first obtained separately for men and women and for younger (age-at-risk <60 years) and older individuals (age-at-risk ≥60 years). The resulting age- and sex-specific HRs and their standard errors were then divided with age- and sex-specific ICCs and were combined by calculating their inverse-variance weighted average.

**Statistical references**

1. Plummer M. Improved estimates of floating absolute risk. Statistics in Medicine. 2004;23(1):93-104.
2. Grambsch PM, Therneau TM. Proportional hazards tests and diagnostics based on weighted residuals. Biometrika. 1994;81(3):515-26.

### Supplementary Tables

#### Table S1: Definitions of coronary heart disease and atherosclerotic cardiovascular disease

| Outcome | ICD-10/OPCS-4 codes | Self-report |
|---|---|---|
| Angina | **Angina pectoris (I20)*** | Angina (UKBFID 6150); Angina (UKBFID 20002) |
| Myocardial infarction | **Acute myocardial infarction (I21)*; Subsequent myocardial infarction (I22)*; Certain current complications following acute myocardial infarction (I23)*** | Heart attack (UKBFID 6150);<br>Heart attack/myocardial infarction (UKBFID 20002) |
| Ischaemic stroke | Cerebral infarction (I63); Stroke, not specified as haemorrhage or infarction (I64); Sequelae of cerebral infarction (I69.3); Sequelae of stroke, not specified as haemorrhage or infarction (I69.4); Sequelae of other and unspecified cerebrovascular diseases (I69.8) | Stroke (UKBFID 6150); Stroke \| Ischaemic stroke (UKBFID 20002) |
| Transient ischaemic attack | Transient cerebral ischaemic attacks and related syndromes (G45) | Transient ischaemic attack (TIA) (UKBFID 20002) |
| Coronary revascularisation | **Revascularisation of wall of heart (K23.4)*; Saphenous vein graft replacement of coronary artery (K40)*; Other autograft replacement of coronary artery (K41)*; Allograft replacement of coronary artery (K42)*; Prosthetic replacement of coronary artery (K43)*; Other replacement of coronary artery (K44)*; Connection of thoracic artery to coronary artery (K45)*; Other bypass of coronary artery (K46)*; Endarterectomy of coronary artery (K47.1)*; Transluminal balloon angioplasty of coronary artery (K49)*; Other therapeutic transluminal operations on coronary artery (K50)*; Percutaneous transluminal balloon angioplasty and insertion of stent into coronary artery (K75)*** | Coronary artery bypass grafts (CABG) \| Triple heart bypass \| Coronary angioplasty (PTCA) +/- stent (UKBFID 20004) |

| Outcome | ICD-10/OPCS-4 codes | Self-report |
|---|---|---|
| Non-coronary revascularisation | Revascularisation procedures including bypass, endarterectomy, endovascular repair, aneurysms repair, embolectomy and percutaneous revascularisation procedures of aorta (L16, L18, L19, L20, L21, L22, L23, L25, L26.1, L26.2, L26.3, L26.5, L26.6, L26.7, L26.8, L26.9, L27, L28), carotid artery (L29, L30, L31.1, L31.3, L31.4, L31.8, L31.9), subclavian artery (L37, L38, L39.1, L39.2, L39.5, L39.8, L39.9), iliac artery (L48, L49, L50, L51, L52, L53, L54.1, L54.2, L54.4, L54.8, L54.9), femoral artery (L56, L57, L58, L59, L60, L62, L63.1, L63.2, L63.5, L63.8, L63.9); Revision of reconstruction of artery (L65); Other therapeutic transluminal operations on artery (L66); Repair of other artery (L68) | Other arterial surgery/revascularisation procedures \| Fem-pop bypass/leg artery bypass \| Leg artery aneurysm repair \| Femoral/popliteal/iliac aneurysm repair \| Aortic aneurysm/repair or stent \| Carotid artery surgery/endarterectomy \| Non-coronary artery angioplasty +/- stent \| Leg artery angioplasty +/- stent \| Carotid artery angioplasty +/- stent (UKBFID 20004) |
| Other atherosclerotic cardiovascular disease | **Other ischaemic heart diseases (I24, I25)***; Atherosclerotic cerebrovascular diseases (I65, I66, I67.2); Atherosclerosis (I70); Aortic and other aneurysms (I71, I72); Peripheral vascular disease (I73.1, I73.8, I73.9, I74, I77.1, K55.1, K55.8, K55.9) | Peripheral vascular disease \| Leg claudication/ intermittent claudication \| Arterial embolism \| Aortic aneurysm \| Aortic aneurysm rupture \| Aortic dissection (UKBFID 20002) |

All codes listed were used to define atherosclerotic disease prior to accelerometer wear, and those marked with an asterisk (*) were used to define incident coronary heart disease. ICD-10, International Statistical Classification of Diseases and Related Health Problems (10th revision); OPCS-4: Office of Population, Censuses and Surveys: Classification of Interventions and Procedures (version 4); UKBFID: UK Biobank Field ID

### Table S2: Descriptions and UK Biobank fields of all reported baseline variables

| Variable | UK Biobank field | Modelling |
|---|---|---|
| **Sociodemographic characteristics** | | |
| Age | 34, 52, 90011 | |
| Sex | 31 | Male, Female |
| Ethnicity | 21000 | Asian, Black, White, Other |
| Geographical region | 54 | London, Wales, NW England, NE England, Yorkshire, West Midlands, East Midlands, SE England, SW England, Scotland |
| Education | 6138 | None of the above, National exams at ages 16-18 years (A levels/AS levels or equivalent, O levels/GCSEs or equivalent, CSEs or equivalent), Vocational qualifications (NVQ or HND or HNC or equivalent, Other professional qualifications eg: nursing, teaching), College or University degree |
| Townsend deprivation index | 22189 | Quintiles |
| **Lifestyle factors** | | |
| Alcohol intake frequency | 1558 | <1, 1-2, 3-4 times/week, Daily drinker |
| Smoking status | 20116 | Never-smoker, Ex-smoker, Current smoker |
| Fruits and vegetables intake | 1289, 1299, 1309 | <4, 4-5.9, 6-7.9, ≥8 servings/day |

| Variable | UK Biobank field | Modelling |
|---|---|---|
| **Physical and blood measurements** | | |
| Body mass index | 21001 | Continuous |
| Systolic blood pressure | 93, 4080 | Continuous |
| Resting heart rate | 102 | Continuous |
| LDL cholesterol | 30780 | Continuous |
| **Self-reported medical history** | | |
| Blood pressure medication | 6153, 6177 | Yes, No |
| Cholesterol medication | 6153, 6177 | Yes, No |
| Long standing illness, disability or infirmity | 2188 | Yes, No |
| Overall health rating | 2178 | Poor, Fair, Good, Excellent |
| Atherosclerotic cardiovascular disease* | 6150, 2000 (category ID) | Yes, No |

* As recorded in hospital records or self-reported (**Table S1**).

**Table S3: Intraclass correlation coefficients (95% CIs) of accelerometer-derived phenotypes of physical activity and sleep, calculated between the main and each repeat measurement, and combined**

| Phenotype | Combined | Repeat 1 | Repeat 2 | Repeat 3 | Repeat 4 |
|---|---|---|---|---|---|
| Overall activity | 0.75 (0.74-0.76) | 0.76 (0.74-0.78) | 0.76 (0.74-0.77) | 0.74 (0.72-0.75) | 0.76 (0.74-0.77) |
| Step count | 0.62 (0.61-0.63) | 0.63 (0.61-0.65) | 0.62 (0.60-0.64) | 0.60 (0.58-0.63) | 0.63 (0.60-0.65) |
| Peak 30-minute cadence | 0.60 (0.59-0.61) | 0.60 (0.57-0.62) | 0.61 (0.59-0.64) | 0.59 (0.56-0.61) | 0.59 (0.57-0.62) |
| Moderate-to-vigorous physical activity | 0.60 (0.59-0.61) | 0.60 (0.57-0.62) | 0.60 (0.58-0.62) | 0.59 (0.56-0.61) | 0.61 (0.58-0.63) |
| Light physical activity | 0.68 (0.67-0.69) | 0.68 (0.65-0.70) | 0.68 (0.66-0.70) | 0.67 (0.65-0.69) | 0.69 (0.67-0.71) |
| Sedentary behaviour | 0.69 (0.68-0.70) | 0.69 (0.67-0.71) | 0.69 (0.67-0.71) | 0.68 (0.66-0.70) | 0.69 (0.67-0.71) |
| Total daily time in bed | 0.61 (0.60-0.62) | 0.61 (0.59-0.64) | 0.62 (0.60-0.64) | 0.60 (0.58-0.63) | 0.60 (0.57-0.62) |
| Overnight sleep duration | 0.61 (0.60-0.62) | 0.61 (0.59-0.64) | 0.62 (0.59-0.64) | 0.61 (0.59-0.63) | 0.59 (0.57-0.62) |
| Sleep efficiency | 0.58 (0.56-0.59) | 0.58 (0.55-0.60) | 0.57 (0.54-0.59) | 0.58 (0.55-0.60) | 0.58 (0.55-0.60) |

Analysis was done in 3138 participants with valid accelerometer data from the main sub-study and at least one valid repeat measurement. Intraclass correlation coefficients were adjusted for season of accelerometer wear (residuals method). Combined estimates were calculated using inverse-variance weighting.

**Table S4: Intraclass correlation coefficients (95% CIs) of accelerometer-derived phenotypes of physical activity and sleep, by subgroups of age and sex**

| Phenotype | Age at main accelerometry sub-study | | Sex | |
|---|---|---|---|---|
| | <60 years (n=1618) | ≥60 years (n=1520) | Men (n=1525) | Women (n=1613) |
| Overall activity | 0.71 (0.69-0.72) | 0.77 (0.76-0.78) | 0.72 (0.71-0.73) | 0.79 (0.77-0.80) |
| Step count | 0.58 (0.56-0.60) | 0.66 (0.65-0.68) | 0.58 (0.57-0.60) | 0.66 (0.64-0.67) |
| Peak 30-minute cadence | 0.54 (0.53-0.56) | 0.63 (0.61-0.64) | 0.58 (0.56-0.59) | 0.61 (0.60-0.63) |
| Moderate-to-vigorous physical activity | 0.58 (0.57-0.60) | 0.61 (0.60-0.63) | 0.57 (0.55-0.59) | 0.61 (0.59-0.63) |
| Light physical activity | 0.67 (0.66-0.69) | 0.68 (0.67-0.70) | 0.65 (0.64-0.67) | 0.66 (0.65-0.68) |
| Sedentary behaviour | 0.68 (0.67-0.70) | 0.69 (0.68-0.71) | 0.65 (0.63-0.66) | 0.69 (0.68-0.71) |
| Total daily time in bed | 0.58 (0.56-0.60) | 0.64 (0.62-0.65) | 0.60 (0.59-0.62) | 0.60 (0.58-0.62) |
| Overnight sleep duration | 0.60 (0.58-0.62) | 0.61 (0.60-0.63) | 0.61 (0.59-0.63) | 0.60 (0.58-0.62) |
| Sleep efficiency | 0.56 (0.54-0.58) | 0.59 (0.57-0.60) | 0.58 (0.56-0.60) | 0.57 (0.55-0.58) |

Analytical sample and notations as in **Table S3**.

**Table S5: Intraclass correlation coefficients (95% CIs) of accelerometer-derived phenotypes of physical activity and sleep, by subgroups of body mass index and self-reported disability**

| Phenotype | Body mass index | | Self-reported disability | |
|---|---|---|---|---|
| | <25 kg/m$^2$ (n=1335) | ≥25 kg/m$^2$ (n=1799) | Healthy (n=2253) | Prior disability (n=827) |
| Overall activity | 0.76 (0.75-0.77) | 0.73 (0.71-0.74) | 0.74 (0.73-0.75) | 0.77 (0.75-0.78) |
| Step count | 0.60 (0.58-0.62) | 0.63 (0.61-0.64) | 0.59 (0.57-0.60) | 0.68 (0.66-0.70) |
| Peak 30-minute cadence | 0.58 (0.56-0.60) | 0.59 (0.58-0.61) | 0.58 (0.56-0.59) | 0.60 (0.58-0.63) |
| Moderate-to-vigorous physical activity | 0.56 (0.54-0.58) | 0.62 (0.60-0.63) | 0.58 (0.57-0.60) | 0.62 (0.60-0.64) |
| Light physical activity | 0.69 (0.67-0.70) | 0.66 (0.65-0.68) | 0.67 (0.66-0.68) | 0.69 (0.67-0.71) |
| Sedentary behaviour | 0.66 (0.65-0.68) | 0.68 (0.67-0.70) | 0.67 (0.66-0.69) | 0.71 (0.69-0.73) |
| Total daily time in bed | 0.60 (0.58-0.62) | 0.60 (0.59-0.62) | 0.61 (0.60-0.63) | 0.60 (0.58-0.62) |
| Overnight sleep duration | 0.61 (0.59-0.63) | 0.60 (0.58-0.62) | 0.61 (0.60-0.63) | 0.60 (0.58-0.62) |
| Sleep efficiency | 0.57 (0.55-0.59) | 0.58 (0.56-0.59) | 0.57 (0.56-0.59) | 0.57 (0.55-0.60) |

Analytical sample and notations in **Table S3**; additionally there were 4 participants with missing data for body mass index and 58 with missing data for self-reported disability.

**Table S6: Accelerometer-derived phenotypes of physical activity and sleep across main and repeat accelerometry sub-studies among 1571 participants with four valid repeat measurements**

| Phenotype | Main accelerometry (n=1571) | Repeat 1 (n=1571) | Repeat 2 (n=1571) | Repeat 3 (n=1571) | Repeat 4 (n=1571) |
|---|---|---|---|---|---|
| Time since main accelerometry sub-study, years | 0 | 3.2 (0.6) | 3.5 (0.6) | 3.7 (0.6) | 4.0 (0.6) |
| Current age, years | 59.6 (9.4) | 62.9 (9.3) | 63.1 (9.3) | 63.4 (9.3) | 63.6 (9.3) |
| Women | 812 (51.7%) | 812 (51.7%) | 812 (51.7%) | 812 (51.7%) | 812 (51.7%) |
| Season | | | | | |
| Spring | 335 (21.3%) | 470 (29.9%) | 1141 (72.6%) | 9 (0.6%) | 0 (0%) |
| Summer | 446 (28.4%) | 0 (0%) | 423 (26.9%) | 1156 (73.6%) | 10 (0.6%) |
| Autumn | 506 (32.2%) | 7 (0.4%) | 0 (0%) | 406 (25.8%) | 1163 (74.0%) |
| Winter | 284 (18.1%) | 1094 (69.6%) | 7 (0.4%) | 0 (0%) | 398 (25.3%) |
| Physical activity phenotypes | | | | | |
| Overall activity, mg | 29.0 (8.7) | 27.2 (9.0) | 28.8 (9.2) | 28.2 (8.9) | 27.1 (8.6) |
| Step count, steps | 9600 (3883) | 9074 (3980) | 9908 (3978) | 9799 (4164) | 9126 (3847) |
| Peak 30-minute cadence, steps/minute | 93.7 (16.8) | 92.3 (18.8) | 93.5 (17.0) | 92.6 (17.0) | 92.5 (17.9) |
| Moderate-to-vigorous physical activity, hours | 0.58 (0.30, 1.02) | 0.51 (0.24, 0.93) | 0.62 (0.32, 1.10) | 0.62 (0.30, 1.09) | 0.54 (0.25, 0.99) |
| Light physical activity, hours | 4.9 (1.6) | 4.8 (1.6) | 5.0 (1.6) | 4.9 (1.6) | 4.8 (1.6) |
| Sedentary behaviour, hours | 10.5 (1.8) | 10.5 (1.8) | 10.5 (1.9) | 10.5 (1.9) | 10.6 (1.8) |
| Sleep phenotypes** | | | | | |
| Total daily time in bed, hours | 7.9 (1.0) | 8.0 (1.0) | 7.8 (1.0) | 7.8 (1.0) | 7.9 (1.0) |
| Overnight sleep duration, hours | 6.8 (0.9) | 6.9 (1.0) | 6.8 (1.0) | 6.8 (1.0) | 6.9 (1.0) |
| Sleep efficiency | 0.86 (0.81, 0.90) | 0.86 (0.81, 0.90) | 0.86 (0.81, 0.91) | 0.87 (0.81, 0.91) | 0.86 (0.81, 0.90) |

Notations as in **Table 2**. *After additional measurement quality-related exclusions, descriptive statistics for sleep phenotypes were calculated from the following number of participants: Main (n=1569), Repeat 1 (n=1519), Repeat 2 (n=1545), Repeat 3 (n=1548) and Repeat 4 (n=1552).

**Table S7: Intraclass correlation coefficients (95% CIs) of accelerometer-derived phenotypes of physical activity and sleep among 1571 participants with four valid repeat measurements**

| Phenotype | Intraclass correlation coefficients |
|---|---|
| Overall activity | 0.76 (0.75-0.77) |
| Step count | 0.63 (0.61-0.65) |
| Peak 30-minute cadence | 0.60 (0.59-0.62) |
| Moderate-to-vigorous physical activity | 0.62 (0.60-0.63) |
| Light physical activity | 0.69 (0.67-0.70) |
| Sedentary behaviour | 0.69 (0.68-0.70) |
| Total daily time in bed | 0.61 (0.59-0.63) |
| Overnight sleep duration | 0.61 (0.59-0.63) |
| Sleep efficiency | 0.57 (0.55-0.59) |

Intraclass correlation coefficients were adjusted for season of accelerometer wear (residuals method) and combined from all four repeat measurements using inverse-variance weighting.

**Table S8: Sensitivity analysis of the association between daily step count and coronary heart disease excluding the first 2 years of follow-up**

| Daily step count at main accelerometry (steps) | Main model | | Excluding first 2 years of follow-up | |
|---|---|---|---|---|
| | No. of events | HR (95% CI) | No. of events | HR (95% CI) |
| ≤ 6388 | 1014 | 1.00 (0.94-1.07) | 795 | 1.00 (0.93-1.07) |
| 6389 - 8250 | 789 | 0.84 (0.78-0.90) | 589 | 0.79 (0.73-0.86) |
| 8251 - 10 059 | 786 | 0.84 (0.78-0.90) | 613 | 0.83 (0.77-0.90) |
| 10 060 - 12 465 | 672 | 0.74 (0.68-0.80) | 521 | 0.72 (0.66-0.79) |
| ≥ 12 466 | 638 | 0.71 (0.65-0.76) | 511 | 0.72 (0.66-0.78) |
| Per 4000 usual steps | 3899 | 0.80 (0.76-0.85) | 3029 | 0.82 (0.76-0.87) |

Analytical sample and methods as in **Figure 3**.

**Table S9: Sensitivity analysis of the association between daily step count and coronary heart disease using age- and sex-specific corrections for regression dilution bias**

| Age at risk | Number of participants | Number of events | Regression dilution ratios | Uncorrected HR (95% CI) per 4000 steps | Corrected HR (95% CI) per 4000 usual steps |
|---|---|---|---|---|---|
| **Men** | | | | | |
| <60 years | 13 267 | 286 | 0.55 | 0.95 (0.84-1.07) | 0.91 (0.74-1.12) |
| ≥60 years | 31 160 | 2115 | 0.62 | 0.90 (0.86-0.95) | 0.85 (0.79-0.92) |
| **Women** | | | | | |
| <60 years | 20 625 | 146 | 0.61 | 0.86 (0.72-1.04) | 0.79 (0.58-1.06) |
| ≥60 years | 43 199 | 1352 | 0.71 | 0.80 (0.75-0.85) | 0.73 (0.67-0.80) |
| **Overall association corrected for regression dilution bias** | | | | | **0.81 (0.76-0.85)** |

Analytical sample and methods as in **Figure 3**.

### Supplementary figures

### Figure S1: Flowchart of UK Biobank repeat accelerometry participants included in the reproducibility analysis

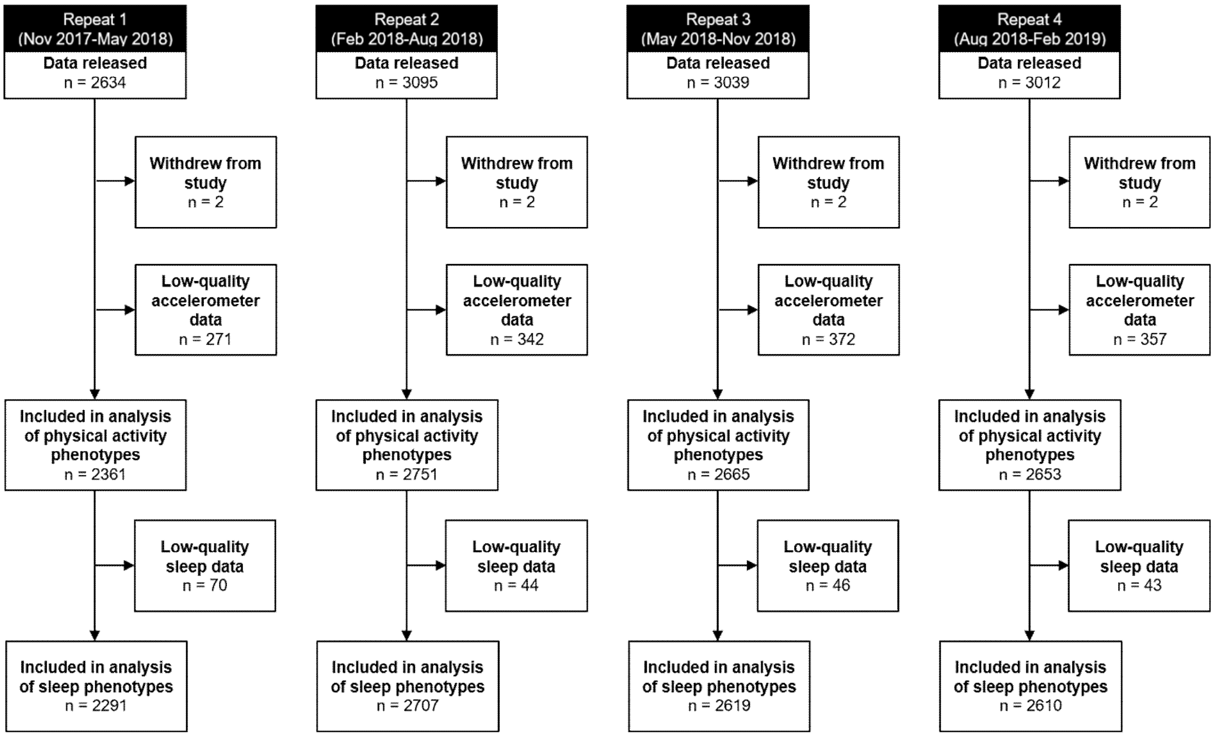

Low-quality accelerometer data was defined as: file failed to be processed, device could not be calibrated, unrealistically high acceleration values (>100mg), insufficient wear time (<3 days of wear or missing data for the same hour across all recorded days in the week). For the investigation of sleep phenotypes, days with less than 22 hours of wear time were additionally excluded.

**Figure S2: Flowchart of UK Biobank main accelerometry participants included in the disease association analysis**

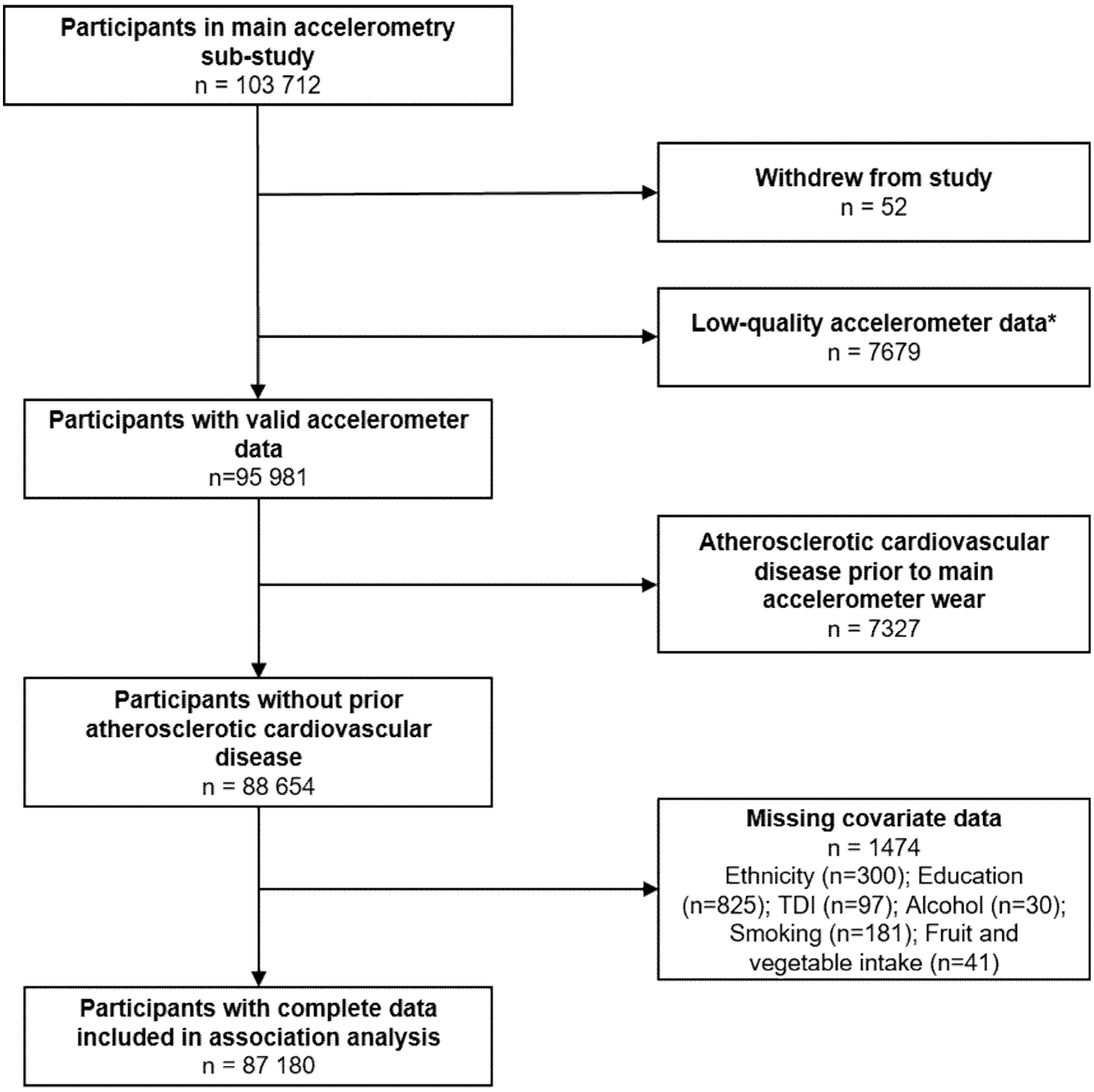

*Low-quality accelerometer data was defined as: file failed to be processed, device could not be calibrated, unrealistically high acceleration values (>100mg), insufficient wear time (<3 days of wear or missing data for the same hour across all recorded days in the week).
